# Effects of joint mobilization combined with acupuncture on pain, physical function, and depression in stroke patients with chronic neuropathic pain: a randomized controlled trial

**DOI:** 10.1101/2023.02.06.23285552

**Authors:** Ji-Eun Lee, Takayuki Akimoto, Ho-Seong Lee

**Author notes:** Corresponding author (HSL).

## Abstract

**Objective:** To investigate the effectiveness of joint mobilization (JM) combined with acupuncture (AC) for treatment pain, physical function and depression in poststroke patients.

**Methods:** A total of 69 poststroke patients were randomly assigned to the JM+AC group (n = 23), the JM group (n = 23), and the control group (n = 23). Patients in the JM+AC group and the JM group received JM for 30 minutes, twice a week for 12 weeks, and the JM+AC group received AC for 30 minutes separately once a week. The control group did not receive JM or AC. Pain (visual analog scale, shoulder pain and disability index, Western Ontario and McMaster universities osteoarthritis index), physical function (range of motion, 10-m walking speed test, functional gait assessment, manual function test, activities of daily living scale, instrumental activities of daily living scale), and depression (center for epidemiologic studies depression scale, Beck depression inventory) were assessed for each patient before and after the 12 weeks of intervention.

**Results:** Pain and physical function were improved significantly in the JM+AC group compared with the JM and control groups. Physical function and depression were improved significantly in the JM+AC and JM groups compared with the control group.

**Conclusion:** The treatment of JM combined with AC improved pain, depression, and physical function of poststroke patients with chronic neuropathic pain in this study. This valuable finding provides empirical evidence for the designing therapeutic interventions and identifying potential therapeutic targets.

## Introduction

Stroke is one of global healthcare problems that is common, serious, and disabling. Motor recovery from stroke is frequently poor or insufficient, with only one-third of poststroke patients regaining dexterity within the first 6 months [1]. Less than 45% of stroke patients are likely to achieve complete functional recovery, and the majority of patients has some residual impairment and an inability to perform activities of daily living (ADL) [2-4]. In addition to motor control and sensory deficits, stroke survivors also develop complications such as pain, spasticity, joint constraint, skin or vascular damage, and chronic neuropathic pain caused by nerve damage, which are the main challenges in poststroke management [5,6].

Shoulder pain experienced by more than 50% of poststroke patients, is a common sequelae [7], interfering with rehabilitation and reducing arm function and ADL [8]. Knee pain is the second most common pain in patients after stroke, and it has been reported to cause motor dysfunction and abnormal gait [9]. The recovery of gait function is a primary goal of rehabilitation, with the majority of poststroke patients suffering from a reduction in gait velocity and independent gait is crucial in carrying out ADL following stroke [10]. Additionally, sensorimotor dysfunction causes limited joint range of motion (ROM) and muscle weakness in the lower extremities of the affected side, leading to difficulties in performing functional activities, such as sit to stand and gait [11,12]. Therefore, it is necessary to reduce pain and increase the joint ROM to improve physical function in poststroke patients. To tackle these problems and to help restore and improve the function of upper and lower limbs, a wide range of treatments has been proposed to therapists, so far.

Mulligan first proposed mobilization-with-movement treatment as a joint mobilization (JM) technique [13]. JM is an orthopedic manipulation therapy used for its mechanical effects to relieve pain, improve mobility, and treat contractures via the third stage of the translatoric movement and the convex–concave rule [14]. According to the convex–concave rule, gliding on a convex joint surface is performed in the opposite direction to the bone’s movement; gliding on a concave joint surface is performed in the same direction as the bone’s movement. When a concave joint surface is distracted, the joint is pulled along the long axis of the bone; alternatively, when a convex joint surface is distracted, the joint surface must be separated [14]. JM has the mechanical effect of disrupting the contracture by direct movement of the joint area, and the arthrokinetic reflex effect of suppressing or facilitating the related muscles is achieved by stimulating the joint receptors [15]. However, there appears to be a limit to how JM alone can reduce pain and restore physical function in poststroke patients.

Acupuncture (AC) is commonly used to treat/manage chronic diseases in Korea [16]. It is recognized to be effective in regulating physiological balance by controlling muscles and acupuncture points or by stimulating the affected area [17]. And AC is effective in reducing pain by stimulating the nerves in stroke patients and help to restore motor function and improve the ADL score [18]. Studies suggest that AC is also effective in improving pain and depression in poststroke patients [19]. Particularly, AC combined with other therapies is more effective in poststroke patients than AC alone [20]. Therefore, it is possible that AC combined with JM is more effective to improve pain, depression, and physical function in poststroke patients. Although there are many studies about effectiveness of JM for poststroke patients, a few studies have examined the effects of JM combined with AC for these patients.

In this work, we examined the changes in pain, physical function, and depression before and after the 12 weeks of intervention of JM combined with AC in poststroke patients. We hypothesized that combining JM with AC would improve physical function more compared with JM alone.

## Methods

### 1. Study design

The study had a random sampling design. Participants were randomized electronically using block sizes of 3 across groups. All investigators and data analysts were blinded until the study and analysis were completed.

Written informed consent was obtained from all subjects and the signed informed consent documents were kept on file. The protocol was approved by the Dankook University ethics committee (IRB number: 2021-05-046) and it adhered to the tenets of the Declaration of Helsinki and its later amendments or comparable ethical standards. The individual in this manuscript has given written informed consent (as outlined in PLOS consent form) to publish these case details.

This clinical trial was registered with the Korean Clinical Research Information Service approved by the WHO. Registration number: KCT0007924. Date registered: 22/11/2022, retrospectively registered. No dropouts or adverse events were reported.

### 2. Participants

This study included 69 adults who were diagnosed with stroke and were enrolled in a public health center and welfare center located in D city, South Korea. This group was further divided into the JM+AC, the JM, and the control groups (n = 23 each).

The inclusion criteria were: diagnosis of stroke ≥ 6 months ago; pain in the shoulders and knees for more than 6 months; and a Korean Mini Mental State Examination score of ≥ 24. The exclusion criteria were: a risk of tumors or infections; a previous history of orthopedic disease or ankle surgery; a previous history of orthopedic disease or shoulder and knee joint surgery; and a length of ≥ 6.3 mm in the line bisection test; taking medications for diseases other than stroke, such as antiplatelet drugs and anticoagulants. The demographics and clinical characteristics of the study patients were summarized at inclusion (Table 1). No significant differences in the pretraining status of the control and experimental groups were noted.

**Table 1.**
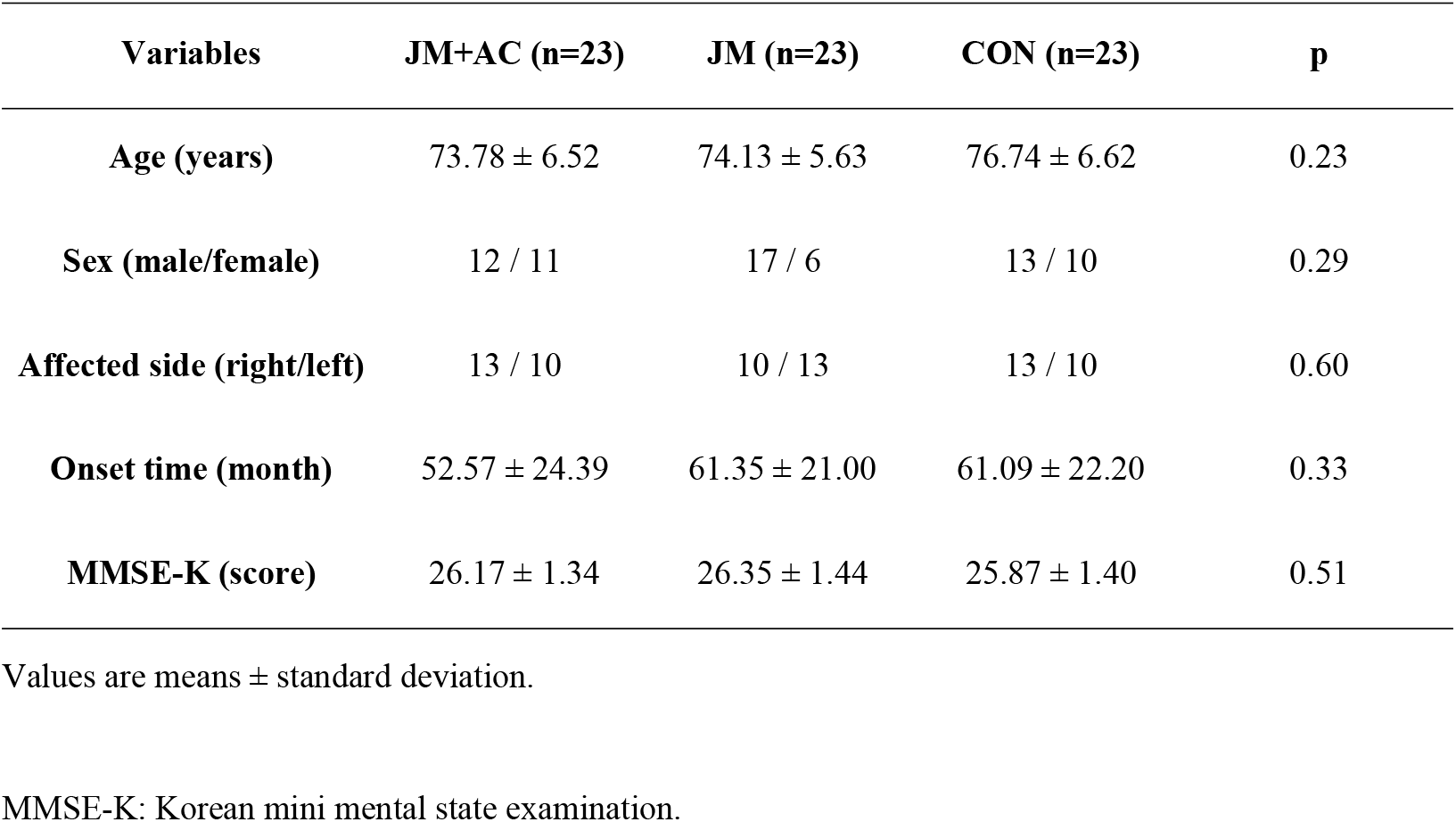
Participant characteristics.

| Variables | JM+AC (n=23) | JM (n=23) | CON (n=23) | p |
| --- | --- | --- | --- | --- |
| Age (years) | 73.78 ± 6.52 | 74.13 ± 5.63 | 76.74 ± 6.62 | 0.23 |
| Sex (male/female) | 12 / 11 | 17 / 6 | 13 / 10 | 0.29 |
| Affected side (right/left) | 13 / 10 | 10 / 13 | 13 / 10 | 0.60 |
| Onset time (month) | 52.57 ± 24.39 | 61.35 ± 21.00 | 61.09 ± 22.20 | 0.33 |
| MMSE-K (score) | 26.17 ± 1.34 | 26.35 ± 1.44 | 25.87 ± 1.40 | 0.51 |

Values are means ± standard deviation.
MMSE-K: Korean mini mental state examination.

### 3. Interventions

#### 3.1. Joint mobilization

For 12 weeks, the JM+AC and JM groups received grades II and III JM for 30 min, twice weekly, to relieve pain in the shoulder and knee joints and to improve the ROM [14]. Shoulder JM was performed on the glenohumeral joint and scapulothoracic joint in the supine and prone positions. Lateral distraction, downward glide, posterior glide, and anterior glide were performed on the glenohumeral joint. Elevation, depression, protraction, retraction, upward rotation, and downward rotation were performed on the scapulothoracic joint. Knee JM was performed on the tibiofemoral and patellofemoral joints in the supine and prone positions. Lower distraction, posterior glide, and anterior glide were performed on the tibiofemoral joint. Downward, lateral, and medial glides were performed on the patellofemoral joint.

#### 3.2 Acupuncture

The JM+AC group received acupuncture and moxibustion treatments around the shoulder and the knee joints for 30 min, once a week for 12 weeks. Acupuncture treatment was performed using disposable sterile stainless-steel needles (0.20 × 30 mm, Dongbang, Korea) for 15 min on average (figure 1). The penetration depth varied according to the site and was 20 mm deep on average. For moxibustion treatment, mini moxibustion (Taegeuk, Korea) was performed for 15 min (fig 1).

**Fig 1.**
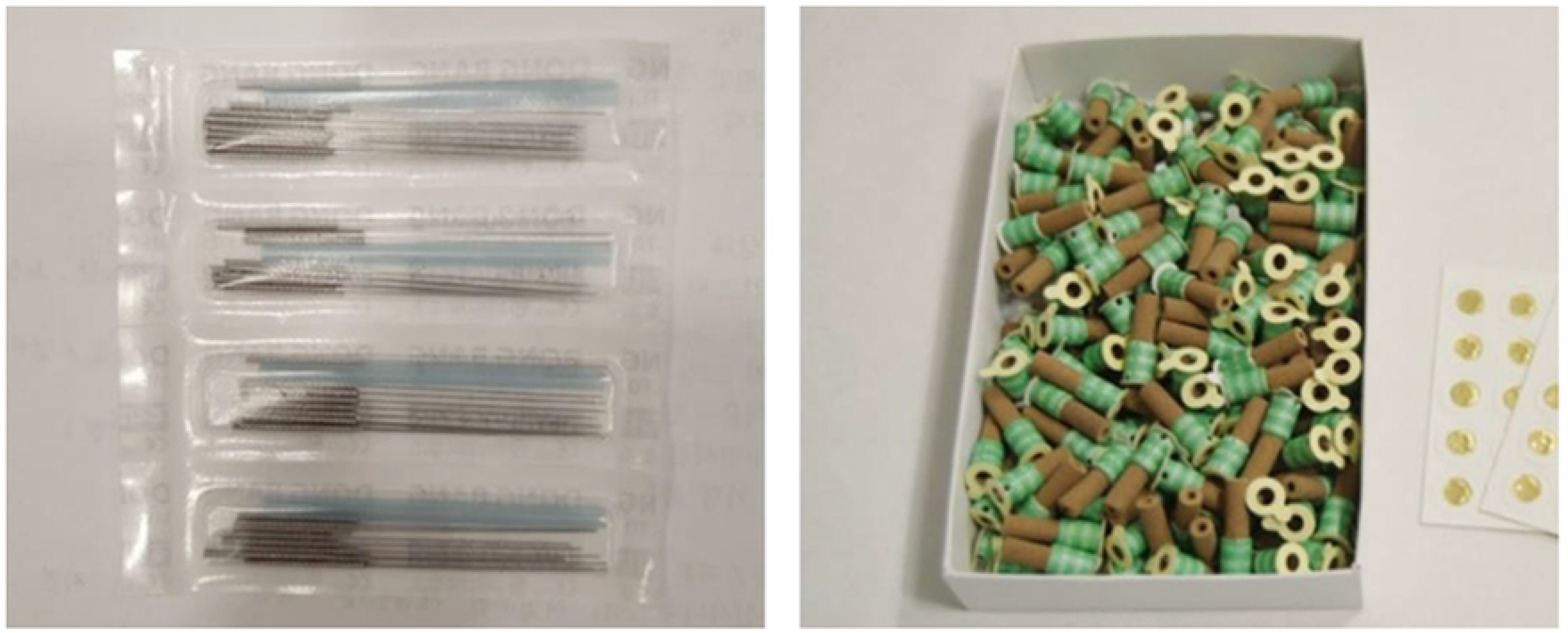
Acupuncture and moxibustion treatment materials. Disposable sterile needles (left picture). Disposable mini moxa rolls (right picture).

### 4. Outcome Measures

#### 4.1 Pain

Pain severity was measured using a visual analog scale (VAS), the shoulder pain and disability index (SPADI), and the Korean Western Ontario McMaster Universities osteoarthritis index (KWOMAC) before and after 12 weeks of the experimental period. The VAS used a 100-mm line on which a pain-free state was 0 and severe pain was 100, and the reliability is 0.99 [21]. SPADI is an evaluation tool for assessing the degree of shoulder pain and disability that comprises 13 evaluation items scored on a scale of 0–10 [22]. A score above 100 (out of 130) indicates more shoulder pain and disability. The reliability is 0.94 [23]. KWOMAC, a Korean version of WOMAC [24], is an evaluation tool for assessing the degree of knee pain and function [25]. It consists of 24 evaluation items scored on a scale of 0–4 points and a total score of 96 points. Higher scores indicate a worse knee condition. The reliability is 0.96 [25].

#### 4.2 Physical function

The range of motion (ROM), 10-meter walk test (10-MWT), functional gait assessment (FGA), manual function test (MFT) for measuring physical function, ADL, and instrumental ADL (IADL) were evaluated before and after 12 weeks of the experimental period.

A goniometer (Saehan Corporation, Korea) was used to measure the ROM of the shoulder and knee joint. Active ROM for shoulder flexion (SF), shoulder extension (SE), shoulder abduction (SAB), shoulder adduction (SAD), and knee flexion (KF) were measured. The measurement was performed three times, and the average value was used. The reliability is 0.89 [26].

The 10-MWT requires the participant to walk independently over the middle 10 m portion on a 14-m path; the time is measured in seconds and the test is repeated three times. The reliability is 0.99 [27].

FGA was developed by Wrisley et al. [28] to evaluate gait and comprises 10 items scored on a scale of 0–3 points and a total score of 30 points. Specific items include walking on level surfaces, changing walking speed, turning the head to the side while walking, moving the head up and down while walking, walking with one foot as an axis, walking around obstacles, gait with a narrow base of support, walking with eyes closed, walking backward, and climbing stairs. The reliability is 0.93 [28].

MFT is a test tool developed to assess the overall condition of arm function in a stroke patient. It consists of eight items, and if subitem inspection is possible, it is measured as 1 point. The reliability is 0.95 [29].

ADL and IADL are tools developed by Won et al. [30] ADL comprising seven items covering dressing, washing the face, bathing, eating, moving, using the toilet, and controlling the bladder, with a total of 21 points. IADL comprises 30 items scored on a scale of 1–3. Ten items cover grooming, chores, preparing meals, washing clothes, going out a short distance, using transport, buying goods, managing money, using a phone, and taking medicine. Higher scores indicate lower dependence in evaluating the independence level in ADL. The reliability is 0.93 [30].

#### 4.3 Depression

Depression was measured using the center for epidemiologic studies depression scale (CES-D) and the Beck depression inventory (BDI) before and after 12 weeks of the experimental period.

CES-D is a self-reported depression evaluation tool [31], comprising 20 evaluation items scored on a scale of 0–3 points. Scores higher than 21 (out of 60) indicate a higher frequency of depressive experiences. The reliability is 0.90 [31].

BDI is a tool for evaluating emotional, cognitive, motivational, and physiological depression, and the Korean version was used [32]. It comprises 21 evaluation items scored on a scale of 0–3 points and a total score of 63 points. The reliability is 0.84 [32].

### 5. Statistical Analysis

The data were analyzed statistically using SPSS 23.0 software (SPSS, Chicago, IL, USA). General characteristics prior to the interventions were analyzed using descriptive statistics. Two-way analysis of variance (2-way ANOVA) with repeated measures was performed to verify the difference between groups and time periods. Multiple comparisons were performed according to the post hoc test (Tukey HSD) when there was a significant difference between groups and time periods. A paired t test was used to compare variations in pain, physical function, and depression before and after experimental period for each group. Statistical significance was reached when p < 0.05.

## Results

### 1. Pain

The changes in pain severity are shown in Table 2. The VAS was not statistically significant between groups, but SPADI decreased significantly in the JM+AC group compared to the JM group and the control group (p < 0.05), and KWOMAC decreased significantly in the JM+AC group compared to the control group (p < 0.05). The VAS, SPADI, and KWOMAC were decreased significantly in the JM+AC and JM groups after 12 weeks of intervention (p < 0.05).

**Table 2.**
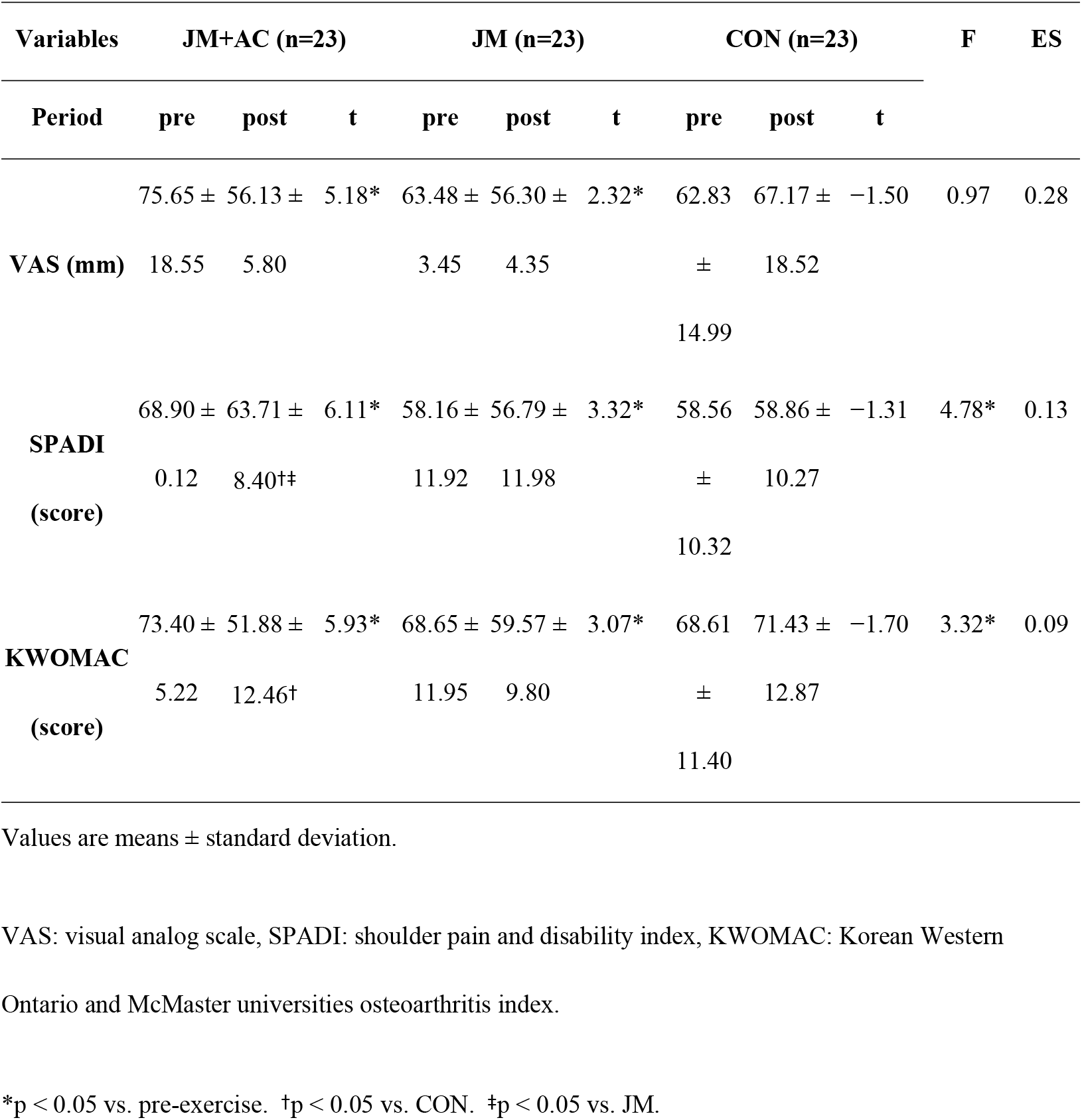
Pain before and after joint mobilization and joint mobilization combined with acupuncture.

### 2. Physical function

The changes in physical functions are shown in Table 3.

**Table 3.**
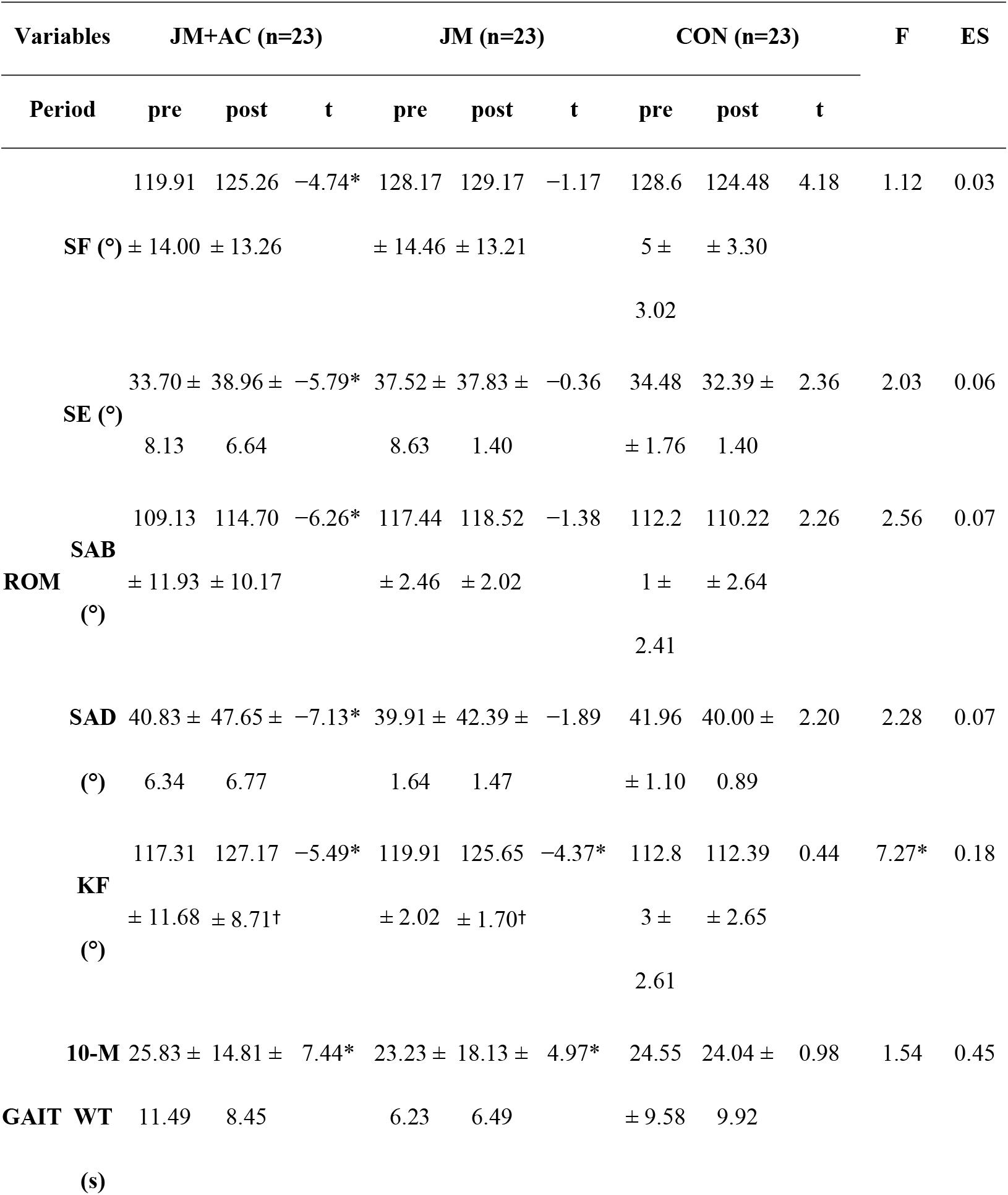
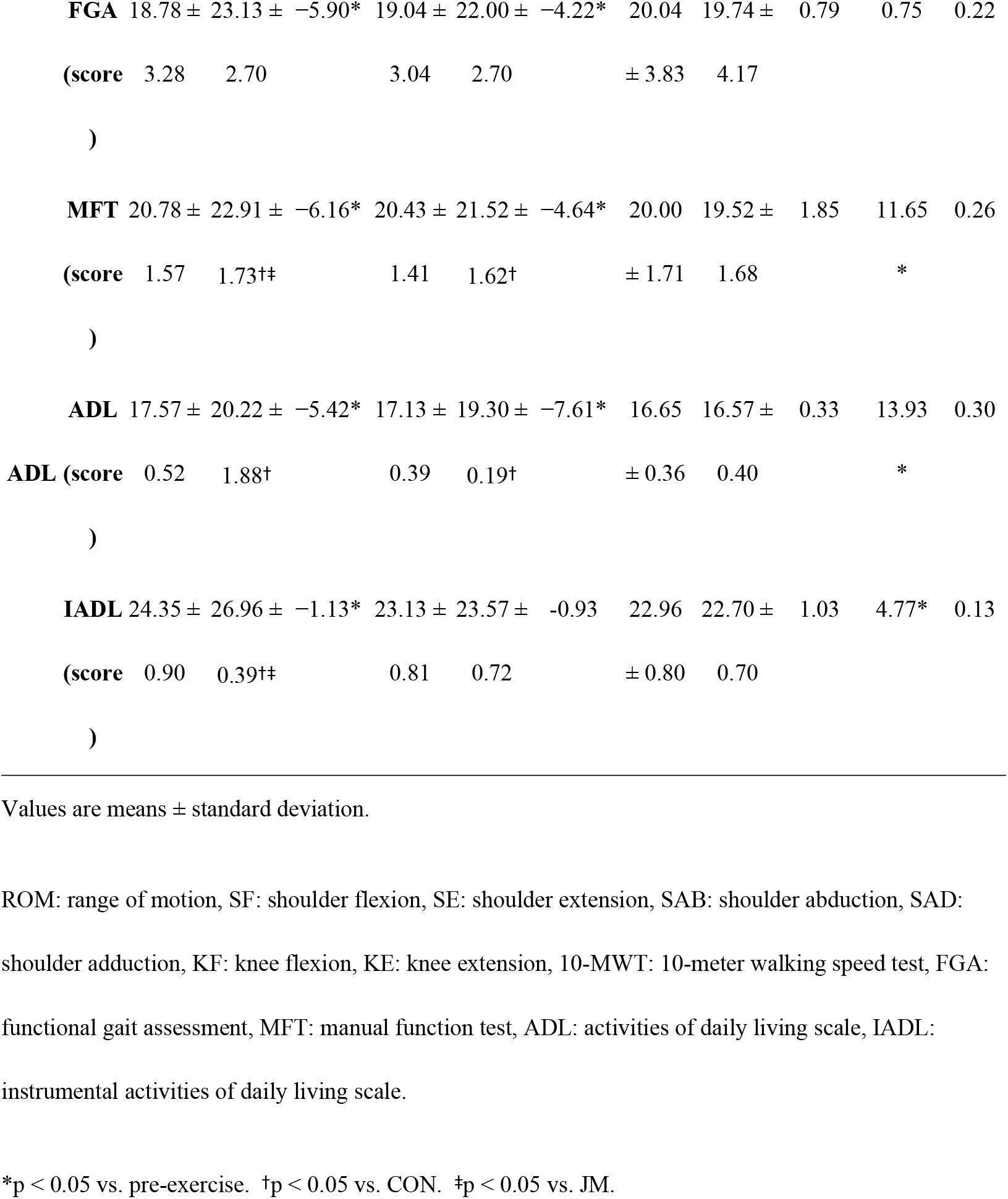
Physical function before and after joint mobilization and joint mobilization combined with acupuncture.

#### 2.1 Range of motion

SF, SE, SAB, and SAD were not statistically significant between groups, but KF increased significantly in the JM+AC and JM groups compared to the control group (p < 0.05). Within the JM+AC group, SF, SE, SAB, and SAD increased significantly (p < 0.05), whereas KF increased significantly in the JM+AC and JM groups after 12 weeks of intervention (p < 0.05).

#### 2.2 Gait

The 10-MWT and FGA were not statistically significant between groups, but 10-MWT and FGA improved significantly in the JM+AC and JM groups after 12 weeks of intervention (p < 0.05).

#### 2.3 Activities of daily living

MFT increased significantly in the JM+AC group compared to the JM group and control groups (p < 0.05), as well as in the JM group compared to the control group (p < 0.05). ADL increased significantly in the JM+AC and JM groups compared to the control group (p < 0.05), and IADL increased significantly in the JM+AC group compared to the JM group and the control group (p < 0.05). Within the JM+AC and JM groups, MFT and ADL increased significantly (p < 0.05), whereas IADL increased significantly in the JM+AC group after 12 weeks of intervention (p < 0.05).

### 3. Depression

The changes in depression are shown in Table 4. CES-D and BDI decreased significantly in the JM+AC and JM groups compared to the control group (p < 0.05), and CES-D and BDI decreased significantly in the JM+AC group after 12 weeks of intervention (p < 0.05).

**Table 4.** Depression before and after joint mobilization and joint mobilization combined with acupuncture.

| Variables | JM+AC (n=23) |  |  | JM (n=23) |  |  | CON (n=23) |  |  | F | ES |
| --- | --- | --- | --- | --- | --- | --- | --- | --- | --- | --- | --- |
| Period | pre | post | t | pre | post | t | pre | post | t |  |  |
| **CES-D** | 26.83 ± 8.63 | 20.30 ± 8.87<sup>†</sup> | 5.24* | 20.48 ± 9.57 | 20.43 ± 8.88 | 0.06 | 27.91 ± 8.96 | 33.00 ± 8.77 | -3.60 | 7.11* | 0.18 |
| **BDI (score)** | 32.48 ± 11.04 | 22.35 ± 7.97<sup>†</sup> | 5.49* | 30.61 ± 12.23 | 29.96 ± 10.99 | 1.19 | 33.70 ± 9.49 | 39.43 ± 9.15 | -3.33 | 5.97* | 0.15 |

Values are means ± standard deviation.
CES-D: center for epidemiologic studies depression scale, BDI: Beck depression inventory.
\* $p < 0.05$ vs. pre-exercise. <sup>†</sup> $p < 0.05$ vs. CON. <sup>‡</sup> $p < 0.05$ vs. JM.

## Discussion

The main finding of this study was that JM combined with AC treatment improved the subjective pain, depression, and physical function of poststroke patients more effectively than JM alone.

Shoulder and knee pain impedes rehabilitation and interferes with daily activities in poststroke patients [7–9]. Kaltenborn et al. [14] indicated that JM was effective in relieves shoulder and knee pain through direct movement of the joints. In AC treatments, acupuncture treatment is reported to increase pain threshold by stimulating the central nervous system, and moxibustion treatment has an analgesic effect through thermal stimulation [18,33]. Additionally, previous studies reported that AC treatment reduced SPADI and KOMAC [34,35]. Taken together, it was confirmed that JM combined with AC treatment had a more positive effect on pain in poststroke patients than JM alone.

Kluding and Santos [36] stated that the JM technique improved movement without changing in joint ROM. Alternatively, Choi et al. [37] reported that AC treatment reduces pain and increases joint flexibility by regenerating the surrounding soft tissue. We found that there was a significant difference in ROM before and after the intervention in the JM+ AC group, but not in the JM group in this study. These results may imply that it is difficult to improve joint ROM with JM alone.

As mentioned in the introduction, gait is an important factor for independent living and social participation. JM showed a positive effect on gait by improving active ROM which is the ROM that can be achieved by patient himself/herself [38], and AC treatment stimulated muscles and nerves to improve gait [39]. Additionally, previous studies stated that acupuncture had a beneficial impact on the energy production of joints during walking [40]. In this study, gait was significantly improved after the intervention in the JM+ AC and JM groups, but there was no difference between groups. Generally, gait necessitates movement of hip, knee, and ankle joints. The lower extremity interventions in this study were limited to the knee joint, which may not have been sufficient to influence group effect comparisons. In the future, it would be prudent to intervene in all joints of lower extremity.

We employed several measurements to access physical function, which is important for daily living. MFT is an assessment of overall arm function [29], SPADI is for shoulder pain and disability [22]. IADL is for evaluation of dressing, toilet using, and movement [30]. It is thought that the reduction of SPADI restored arm function in poststroke patients and affected MFT and IADL. Previous research has shown that acupuncture has little effect on improving the lifestyle of stroke patients [41], but it is believed that it had a positive effect on IADL in this study because joint mobilization was combined with acupuncture. Arm function and daily life activities of the patients in the JM+ AC group might be improved through the increase in muscle activity induced by JM and the recovery of neurological damage by AC treatment.

Depression is associated with pain in poststroke patients [7], and depression in these patients are associated with mortality, suicide risk, and quality of life [42–44]. Thus, depression must be verified in rehabilitation process for functional recovery after stroke, in general. The CES-D and BDI scores in the JM+ AC group were significantly decreased after 12 weeks of the intervention, but not in the JM group. This is most likely due to AC treatment to alleviate pain and improve ADL. A previous study showed that JM improved depression by reducing stress factors through improvement of unstable posture [45]. Furthermore, it has been reported that acupuncture treatment was effective in suppressing changes in the autonomic nervous system caused by mental stress [46]. It has also been reported that moxibustion treatment relaxed and stabilized the body through its thermal action [18]. Thus, it is considered that acupuncture and moxibustion treatment of AC reduced depression through autonomic nerve inhibition and thermal stimulation.

### Limitation

There are several limitations in our study. The major part of subjects in this study was older farmers living in rural areas, and it is not uncertain whether the results of this study can be applied to other elderly population living in urban areas. Additionally, it might be better if we assessed the effect of the treatment more frequently because the results of this study were obtained before and after the 12 weeks of intervention period.

## Conclusion

Improvements in pain, depression, and physical function of poststroke patients in the JM+ AC group were significantly greater than those in the JM and the control groups, suggesting JM combined with AC was more effective in improving not only pain but also the ability to physical function. JM combined with AC may also be helpful for poststroke patients who have depression and decreased physical function due to chronic neuropathic pain. Future studies should reveal the physiological assessment of muscle strength and the effect of JM combined with AC on muscle tone based on changes in physical function in stroke patients with chronic neuropathic pain.

## Data Availability

All data files are available from the public repository Figshare database (https://doi.org/10.6084/m9.figshare.21789092).

## Author Contributions

Conceived and designed the experiments: JEL. Performed the experiments: JEL. Analyzed the data: JEL, TA. Wrote the paper: JEL, TA. Revised paper: TA, HSL.

## Conflicts of Interest

The authors declare that there are no conflicts of interest regarding the publication of this paper.

## Acknowledgments

We appreciate Kim Yeon-oh and Choi Jong-bin of the Dangjin City Public Health Center for their assistance with this experiment.

## Data Availability Statement

Access to data was prepared using the following data set and is available at the free, public repository Figshare (https://doi.org/10.6084/m9.figshare.21789092).

## Funding

This research did not receive any specific grant from funding agencies in the public, commercial, or not-for-profit sectors.

## Supporting Information

**S1 Fig. Acupuncture and moxibustion treatment materials**. Disposable sterile needles (left picture). Disposable mini moxa rolls (right picture).

**S1 Table. Participant characteristics**. MMSE-K: Korean mini mental state examination.

**S2 Table. Pain before and after joint mobilization and joint mobilization combined with acupuncture**. VAS: visual analog scale, SPADI: shoulder pain and disability index, KWOMAC: Korean Western Ontario and McMaster universities osteoarthritis index.

**S3 Table. Physical function before and after joint mobilization and joint mobilization combined with acupuncture**. ROM: range of motion, SF: shoulder flexion, SE: shoulder extension, SAB: shoulder abduction, SAD: shoulder adduction, KF: knee flexion, KE: knee extension, 10-MWT: 10-meter walking speed test, FGA: functional gait assessment, MFT: manual function test, ADL: activities of daily living scale, IADL: instrumental activities of daily living scale.

**S4 Table. Depression before and after joint mobilization and joint mobilization**

**combined with acupuncture**. CES-D: center for epidemiologic studies depression scale, BDI: Beck depression inventory.

**S1 Checklist. CONSORT Checklist**. Checklist of information to include when reporting a randomized trial.

**S2 Checklist. CONSORT Flow Diagram**. Flow diagram.

## Notes

### Competing Interest Statement

The authors have declared that no competing interests exist.

### Clinical Trial

This clinical trial was registered with the Korean Clinical Research Information Service (CRIS) approved by the WHO. Registration number: KCT0007924. Date registered: 22/11/2022, retrospectively registered. No dropouts or adverse events were reported.

### Clinical Protocols

https://cris.nih.go.kr/cris/search/detailSearch.do/23449

### Funding Statement

The authors received no specific funding for this work.

### Author Declarations

This study complied with the guidelines of the Helsinki Declaration and was approved by the Institutional Review Board of Dankook University (DKU No: 2021-05-046).

